# Salivary SARS-CoV-2 antigen rapid detection: a prospective cohort study

**DOI:** 10.1101/2020.12.24.20248825

**Authors:** Daniela Basso, Ada Aita, Andrea Padoan, Chiara Cosma, Filippo Navaglia, Stefania Moz, Nicole Contran, Carlo-Federico Zambon, Anna Maria Cattelan, Mario Plebani

## Abstract

**Background:** SARS-CoV-2 quick testing and reporting are now considered relevant for the containment of new pandemic waves. Antigen testing in self-collected saliva might be useful. We compared the diagnostic performance of salivary and naso-pharyngeal swab (NPS) SARS-CoV-2 antigen detection by a rapid chemiluminescent assay (CLEIA) and two different point-of-care (POC) immunochromatographic assays, with that of molecular testing.

**Methods:** 234 patients were prospectively enrolled. Paired self-collected saliva (Salivette) and NPS were obtained to perform rRT-PCR, chemiluminescent (Lumipulse G) and POC (NPS: Fujirebio and Abbott; saliva: Fujirebio) for SARS-CoV-2 antigen detection.

**Results:** The overall agreement between NPS and saliva rRT-PCR was 78.7%, reaching 91.7% at the first week from symptoms onset. SARS-CoV-2 CLEIA antigen was highly accurate in distinguishing between positive and negative NPS (ROC-AUC=0.939, 95%CI:0.903-0.977), with 81.6% sensitivity and 93.8% specificity. This assay on saliva had an overall good accuracy (ROC-AUC=0.805, 95%CI:0.740-0.870), reaching the optimal value within 7 days from symptom onset (Sensitivity: 72%; Specificity: 97%). POC antigen in saliva had a very limited sensitivity (13%), performing better in NPS (Sensitivity: 48% and 66%; Specificity: 100% and 99% for Espline and Abbott respectively), depending on viral loads.

**Conclusions:** Self-collected saliva is a valid alternative to NPS for SARS-CoV-2 detection not only by molecular, but also by CLEIA antigen testing, for which the highest diagnostic accuracy was achieved in the first week from symptom onset. Saliva is not suitable for POC, although the accuracy of these tests appears satisfactory for NPS with high viral load.

## Introduction

Saliva testing for SARS-CoV-2, one of the strategies for COVID-19 diagnosis and monitoring, is advocated mainly for screening asymptomatic subjects in order to rapidly detect and isolate infected individuals and their contacts, thus limiting viral spread and containing further waves of the pandemic (1-6). Although the molecular detection of SARS-CoV-2 RNA in naso-pharyngeal swabs (NPS) is considered the “gold standard” technique for identifying symptomatic or asymptomatic individuals (7), it has limitations in both the analytical and the healthcare settings. From the analytical viewpoint, it is widely agreed that the sensitivity of rRT-PCR of NPS ranges from 70 to 90% (5,8), reaching values around 50% after the first two weeks of disease (9,10). Therefore, COVID-19 disease cannot be ruled out when a NPS result is negative, but the patient has clinical symptoms, and biochemical data and radiological findings that evidence a clinical scenario typical of the disease (11). In this context anti-SARS-CoV-2 antibodies should also be taken into account (12). From the healthcare organizational viewpoint, NPS testing calls for the involvement of healthcare workers and services for sample collection. This pre-requisite might be promptly met by the healthcare system when the demand is low, but not necessarily when it is high, as occurs in a pandemic. Any delay in testing puts individuals without a diagnosis at risk, consequently exposing the community to viral contagion.

Saliva testing might not only have the advantage of relieving health care resources, but also of reducing hazard exposure to healthcare workers during sampling; it might also limit the risk of viral spread incurred when numerous individuals queue for a long time waiting for testing, since saliva can be self-collected at home. However, none of these advantages support the use of saliva testing if its results are not as reliable as those of NPS. When rRT-PCR is used for SARS-CoV-2 testing, the reliability of saliva testing is reportedly equal to, or even higher than, NPS (5,10,13,14), although some studies report contradictory findings (i.e. saliva as less sensitive) (15). This discrepancy might depend on the salivary viral load kinetic, the highest load occurring in the first week of symptom onset, followed by a progressive decline during the course of the disease (3,9,14). Moreover, the viral load estimated by the threshold cycle (Ct) has been found to be higher in saliva than in NPS especially in pre-symptomatic individuals, thus supporting the view that these subjects increase viral spread (16).

Saliva testing by rRT-PCR is reliable, but time consuming, calling for dedicated laboratory equipment for nucleic acid extraction and amplification, and personnel trained in molecular techniques (17). These requirements, which might not be fulfilled by all laboratories, compromise the advantage of using the saliva sample. In front of a safe and rapid collection procedure, the overall testing process remains long not only because molecular testing takes time, but also because molecular laboratories might be limited in number, especially in low resource countries, thus increasing the turnaround time due to sample transportation and processing. In order to speed up testing while maximizing the number of tested individuals, the search for SARS-Co-2 antigens rather than RNA, by immunometric techniques is now emerging, including point-of-care (POC) rapid immunochromatographic assays based on lateral flow technology (18). The market now offers a number of SARS-CoV-2 antigen detection immunometric assays, which are high throughput but require laboratory instrumentation, and ultra-rapid POC devices suggested for use in medical cabinets by general practitioners and nurses. Since they are simple to use, these devices are also considered potentially employable in auto-testing. However, simplicity is not synonymous with accuracy. Assays designed to identify SARS-CoV-2 antigens have been investigated using NPS samples with a reported sensitivity ranging from 0 to 94% for rapid immunochromatographic assays (19), and from 85 to 100% (depending on the viral load) and a specificity above 99% for antigen levels measured by automated chemiluminescent assay (20). No exhaustive data are present in the literature on antigen detection using saliva, an approach that might maximize an effective and timely COVID-19 diagnosis, encompassing the advantages of a) saliva self-collection and b) rapid viral protein detection, thus making wide-scale screening possible in many parts of the world.

The aim of this prospective study was to compare in saliva and NPS the diagnostic accuracy of molecular testing with SARS-CoV-2 antigen detection by a rapid chemiluminescent assay and two different point of care ultra-rapid immunochromatographic assays.

## Materials and methods

### Patients and samples

A total of 234 subjects were enrolled between 1 August and 30 November 2020. One hundred thirty-eight (52 females, 86 males, mean age±SD: 56±17 years) were COVID-19 inpatients, and 96 (47 females, 49 males, mean age±SD: 42±15 years) were outpatients screened for suspected SARS-CoV-2 (i.e. contact with a SARS-CoV-2 positive subject or with typical symptoms). After obtaining giving fully informed consent in writing (Local Ethic Committee Nr. 27444), patients were asked to collect a morning saliva sample (Salivette device, SARSTEDT AG & Co, Nümbrecht, Germany).

Saliva was self-collected by the Salivette device (SARSTEDT AG & Co, Nümbrecht, Germany), the cotton swab being chewed for at least one minute to stimulate salivation. In order to obtain clear saliva, the Salivette device was centrifuged at 4000 g for 5 minutes within 3 hours of collection. An aliquot of saliva (200 μL) was used for rRT-PCR, and another aliquot (150 μL) for chemiluminescence (CLEIA) SARS-CoV-2 antigen determination. A third saliva aliquot (about 100 μL) was used for SARS-CoV-2 antigen testing with the immunochromatographic ESPLINE rapid test (Fujirebio, Tokjo, Japan) following the manufacturer’s instructions for this sample type.

After saliva sampling, trained nurses collected three NPS from each patient. One was collected using the ∑-VCM (Medical Wire & Equipment, UK) and used for SARS-CoV-2 molecular and CLEIA antigen testing. The second and third swabs were used for SARS-CoV-2 antigen testing by the immunochromatographic ESPLINE rapid test and COVID-19 Ag Rapid Test (ABBOTT, Chicago, Illinois, USA) at the bedside, following the manufacturers’ instructions. The former test requires 30 minutes while the latter 15 minutes for reading. A subset of 32 inpatients repeated saliva collection and testing 7 days after enrollment and, among them, 23 repeated also NPS. Clinical data on each inpatient were also retrieved from the hospital information system (HIS).

### Repeatability and intermediate precision evaluation

Precision estimation was performed on CLEIA assays using three NPS and three saliva pools made up of ten different samples each. Triplicate aliquot measurements were performed over three consecutive days. For estimating precision we used analysis of variance following the CLSI EP15-A3 protocol (CLSI. User Verification of Precision and Estimation of Bias; Approved Guideline— Third Edition. CLSI EP15-A3. Wayne, PA: Clinical and Laboratory Standards Institute, 2014). Repeatability and within-laboratory precision were in accordance with the repeatability and intermediate precision conditions specified in the international vocabulary of metrology (VIM, JCGM 100:2012) for precision estimation within a three-day period.

### Laboratory testing

All molecular and CLEIA antigen testing in both saliva and NPS were performed in parallel within 3 hours from collection. For molecular testing, saliva and swab samples were mixed (2:1, v:v) with Nuclisens^®^ easyMAG^®^ Extraction buffer 1 immediately before extraction of RNA using an automated platform (Magna Pure 96 Instrument, Roche Diagnostics, USA), and then used for Orf1ab, N and S SARS-CoV-2 genes rRT-PCR, by means of the diagnostic system TaqPath COVID-19 RT-PCR kit (Applied Biosystems, USA), performed by QuantStudio™ 5 Real-Time PCR Systems (Applied Biosystems, USA). RNaseP was also separately analyzed by QuantStudio™ 5 RealTime PCR Systems (Applied Biosystems, USA) as described elsewhere [21]. In the absence of RNaseP target amplification, the analysis was considered invalid, and this criterion was used to decide on repeat testing. The threshold cycle (Ct) of SARS-CoV-2 genes Orf1ab, N and S, and of RNaseP was obtained after standardization of the rRT-PCR instrument’s software settings as follows: baseline calculated in the cycle range 3 to 15; fixed threshold, 0.1. The Ct of each analysis was considered for data comparison. In each analytical set, two negative and two positive controls were always run in parallel with the subject’s samples. The positive controls (IQC) were RNA, obtained from positive samples stored at −80 °C for a maximum of one week. Saliva and NPS samples were considered positive when at least two of three targets had an amplification plot with a Ct value of < 40.

Chemiluminescence immunoassay (CLEIA) was performed using an LUMIPULSE SARS-CoV-2 Ag kit on a LUMIPULSE G1200 automated analyzer (Fujirebio, Tokjo, Japan), following the manufacturer’s instructions. After the first result, available in 30 minutes, the diagnostic system provides 120 results per hour. ESPLINE rapid test and COVID-19 Ag Rapid Test (ABBOTT, Chicago, Illinois, USA) were the two evaluated POC.

### Statistical analysis of data

The statistical analysis of data was made with Stata software ver. 13.1 (Lakeway drive, TX, US). Descriptive statistics included mean and standard deviation or median and interquartile ranges (25^th^ and 75^th^ percentile) where appropriate. The following tests were applied: Wilcoxon rank test, Kruskal-Wallis rank test, Student’s t test, Fisher’s exact test, and multiple linear regression. Bonferroni’s adjustment was used for multiple comparisons. Non-parametric ROC analyses were used to estimate the area under the ROC curve. Sensitivity and specificity were estimated by means of the user community package “diagt”.

## Results

### Clinical data

The clinical characteristics of the 138 inpatients are shown in Table 1. While no significant differences were found between in- and outpatients (n=96) for gender distribution (Fisher’s exact test: p=0.106), the mean age of outpatients was significantly lower than that of inpatients (Student’s t test for unpaired data: t=6.51, p<0.001).

**Table 1.**
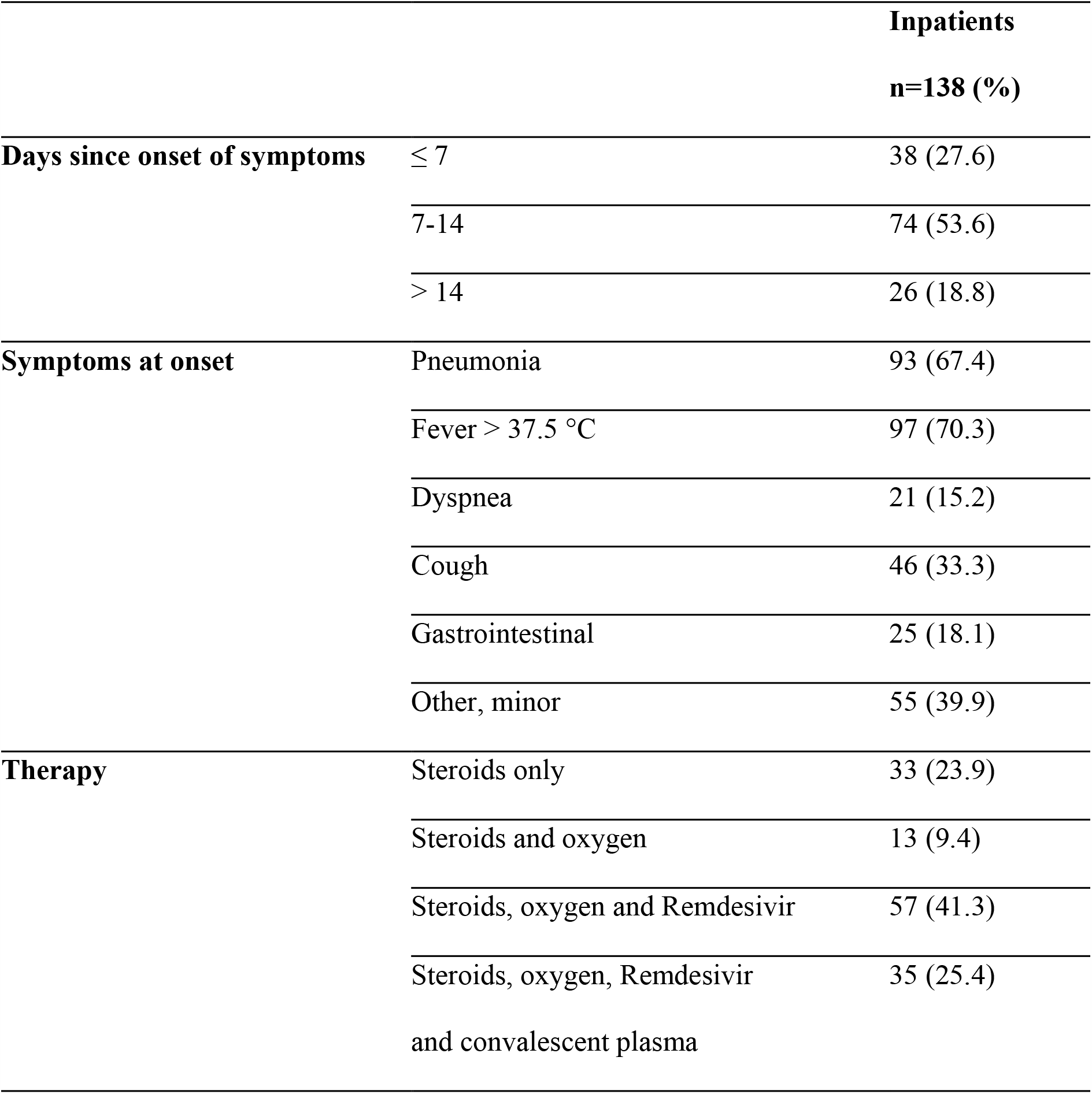
Clinical characteristics of inpatients.

### NPS and saliva molecular testing

At enrollment, NPS rRT-PCR results were positive among 84/138 (60.9%) inpatients and 3/96 (3.1%) outpatients, while saliva in 67/127 (52.8%) inpatients and in 4/96 (4.2%) outpatients. Positive NPS and saliva were more frequent among inpatients with a time lapse from symptoms of less than 7 days (37/38, 97.4% for NPS; 32/36, 88.9% for saliva), with respect to those with 7 to 14 days’ (40/74, 54.1% for NPS; 31/69, 44.9% for saliva) and those with more than 14 days’ (7/26, 26.9% for NPS, 4/22, 18.2% for saliva) (Fisher’s exact test: p<0.001 for both NPS and saliva). Accordingly, the lowest median Ct values of NPS and saliva were found among inpatients with a time lapse from symptoms of less than 7 days (Table 2).

**Table 2.** Median values with interquartile range of the Ct values obtained in NPS and saliva for the three SARS-CoV-2 analyzed genes. Patients were subdivided on the basis of the time lapse between onset of symptoms and enrollment. P/T= number of positive samples out of the total number of samples examined. Each sample was obtained from one individual patient.

| NPS |  |  |  | Saliva |  |  |  |
| --- | --- | --- | --- | --- | --- | --- | --- |
| Time | Orf1ab | Gene N | Gene S | Time | Orf1ab | Gene N | Gene S |
| lapse | Median | Median | Median | lapse | Median | Median | Median |
| (P/T) | IQR | IQR | IQR | (P/T) | IQR | IQR | IQR |
| < 7 days | (n=28) | (n=28) | (n=25) | < 7 days | (n=32) | (n=29) | (n=26) |
| (28/37) | 25.1* | 25.6* | 26.3* | (32/36) | 28.7 | 28.6 | 29.9 |
|  | 19.8-29.0 | 19.9-29.1 | 22.7-29.8 |  | 24.2-32.8 | 23.4-32.9 | 25.4-33.6 |
| 7-14 days | (n=35) | (n=35) | (n=32) | 7-14 days | (n=26) | (n=26) | (n=24) |
| (35/40) | 30.5 | 30.7 | 31.3 | (31/69) | 31.3 | 31.2 | 30.9 |
|  | 26.2-33.1 | 26.5-32.6 | 27.2-33.5 |  | 27.3-33.7 | 28.1-34.1 | 28.4-35.4 |
| > 14 days | (n=4) | (n=4) | (n=6) | > 14 days | (n= 3) | (n=4) | (n=3) |
| (4/7) | 31.4 | 31.5 | 32.8 | (4/22) | 33.2 | 33.0 | 34.0 |
|  | 30.2-34.7 | 30.2-32.7 | 31.7-34.0 |  | 31.0-38.2 | 32.3-34.6 | 33.5-36.2 |
| Kruskal- | $\chi^2=12.6$ | $\chi^2=12.3$ | $\chi^2=11.1$ | | $\chi^2=3.93$ | $\chi^2=5.51$ | $\chi^2=4.71$ |
| Wallis | p=0.002 | p=0.002 | p=0.004 |  | p=0.139 | p=0.064 | p=0.095 |
| test |  |  |  |  |  |  |  |

Among the 96 outpatients, four had positive findings at NPS and/or saliva testing (Table 3). The patient who was negative at NPS but positive at saliva (n. 4), repeated NPS two and ten days after enrollment, being positive in both cases.

**Table 3.**
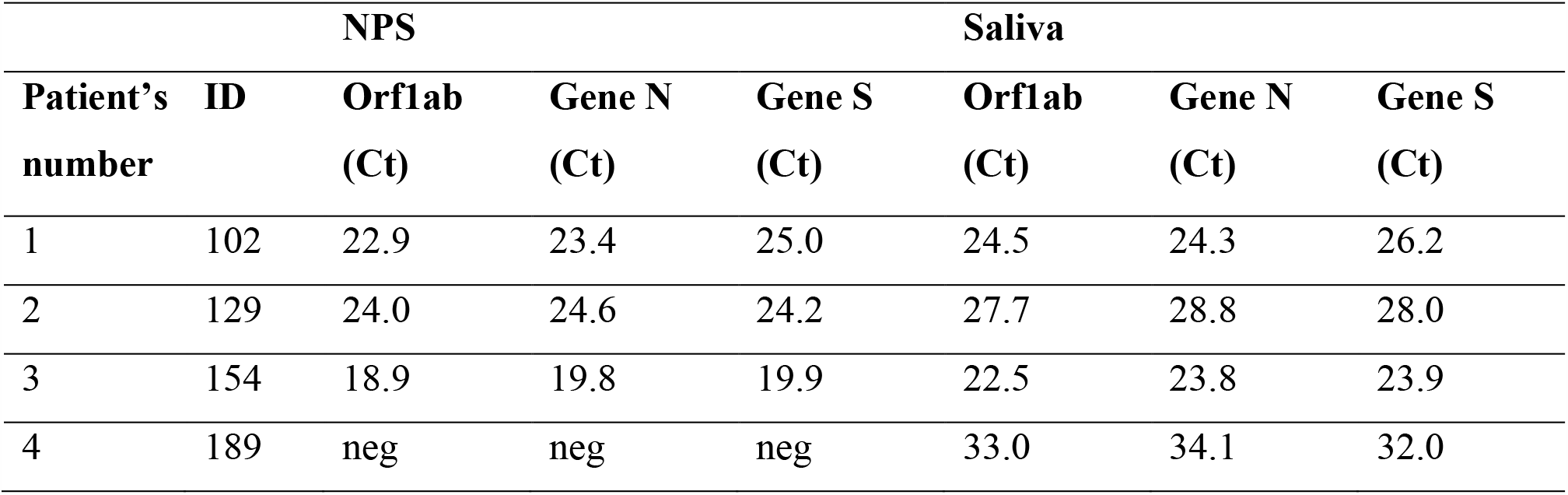
Individual positive rRT-PCR results at NPS and saliva found among outpatients.

On considering the overall inpatient population, agreement of 78.7% was found for rRT-PCR among paired NPS and saliva samples (n=127, Cohen k=0.569, SE=0.086, p<0.001). Agreement was 91.67% in patients tested less than 7 days after onset of symptoms (n=36, Cohen k=0.372, SE=0.130, p<0.001), 71.0% in those tested after 7 to 14 days (n=69, Cohen k=0.427, SE=0.117, p<0.001) and 81.8% in those after more than 14 days (n=22, Cohen k=0.488, SE=0.206, p=0.009).

### NPS and saliva antigen testing

Figure 1 shows the individual CLEIA antigen levels measured in NPS and saliva after subdividing patients on the basis of the time lapse between symptom onset and testing, data on analytical reproducibility being reported in Table 4. SARS-CoV-2 antigen measured in NPS enabled distinction between positive and negative swabs classified on the basis of rRT-PCR with a high diagnostic accuracy (area under the ROC curve=0.939, 95% CI: 0.903-0.977). Based on the threshold reported by the manufacturer (1.34 ng/L), the overall sensitivity and specificity were 81.6% (95% CI: 71.0-89.5%) and 93.8 (95% CI: 86.2-98.0%), respectively, with a positive likelihood ratio (LR) and negative LR of 13.2 (95% CI: 5.62-31.1) and 0.19 (95% CI: 0.12-0.32), respectively.

**Table 4.** Repeatability and intermediate precision of SARS-CoV-2 antigen determination by CLEIA using NPS or saliva. For all levels, a pool of ten individual samples were prepared and tested three times daily for three consecutive days.

| **Matrix type** | **ng/L level** | **% Repeatability** | **% Intermediate precision** |
| --- | --- | --- | --- |
| NPS | 0.60 | 5.7 | 33.4 |
| NPS | 1.22 | 8.8 | 13.1 |
| NPS | 12.00 | 2.8 | 6.6 |
| Saliva | 0.1 | 68 | 72.5 |
| Saliva | 11.8 | 4.5 | 4.5 |
| Saliva | 2913 | 5.4 | 14.5 |

**Figure 1.**
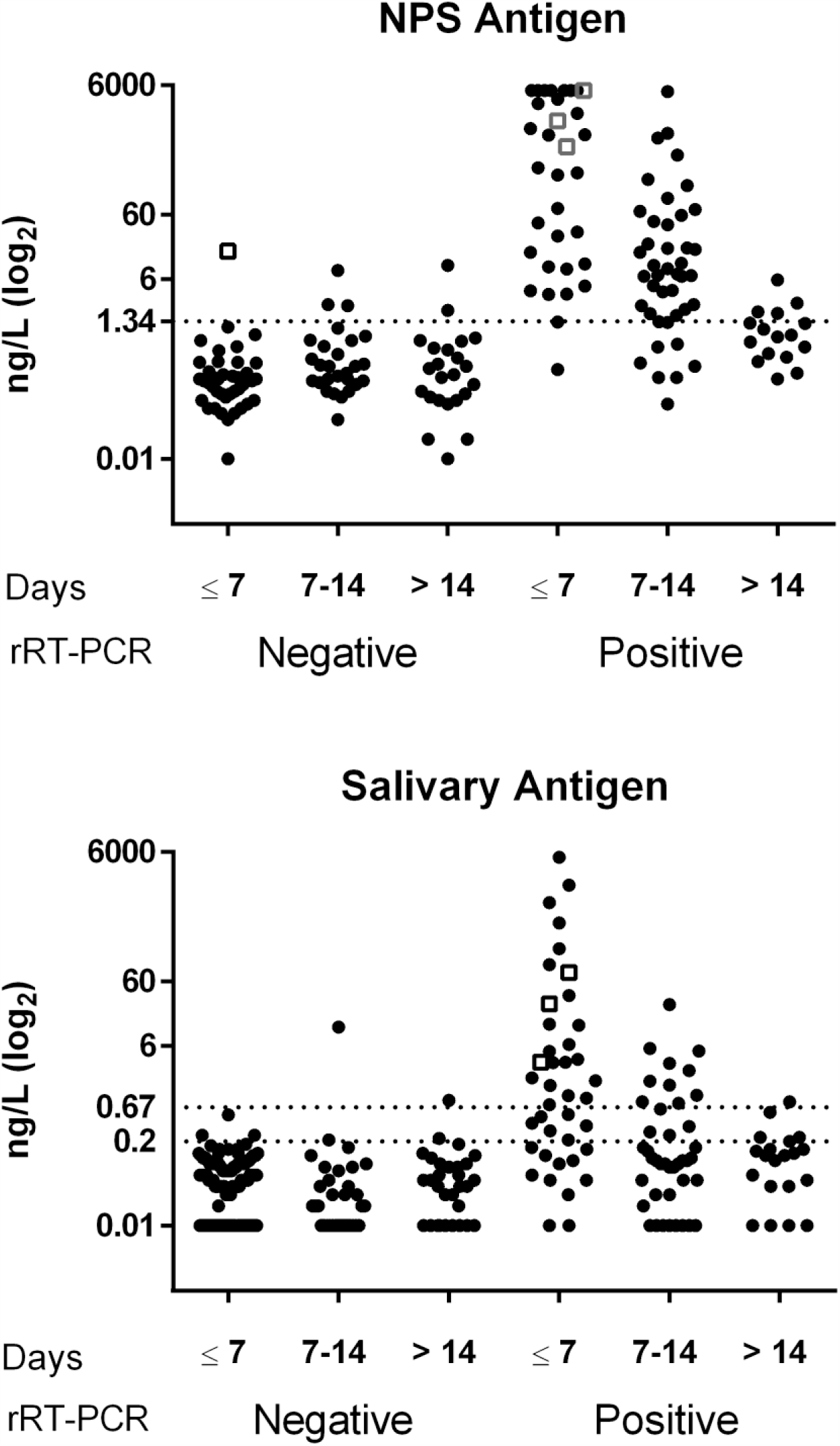
SARS-CoV-2 antigen in NPS and saliva. The antigen was assayed by CLEIA in subjects classified as negative or positive on the basis of NPS rRT-PCR and subdivided on the basis of the time lapse between symptom onset and testing (Days). The upper graph shows the results obtained in NPS samples. The dotted line (1.34 ng/L) is the cut-off recommended by the manufacturer. The lower graph shows the results obtained in salivary samples. The dotted lines are the cut-off (0.67 ng/L) and the limit of detection (0.2 ng/L) recommended by the manufacturer. In both graphs, patients enrolled within 7 days and classified as negative are all but one (open square) outpatients. Among rRT-PCR positive results, open squares represent the three outpatients who were found to be positive.

Salivary SARS-CoV-2 antigen allowed to distinguish between positive and negative samples classified on the basis of NPS rRT-PCR with a good diagnostic accuracy (area under the ROC curve=0.805, 95% CI: 0.740-0.870) (Table 5). For salivary antigen, the manufacturer’s suggested cut-off is 0.67 ng/L and the limit of quantification, 0.2 ng/L. With these two thresholds, sensitivity, specificity, positive and negative LR were calculated considering the patients overall and after subdividing them on the basis of duration of symptoms (Table 6).

**Table 5.** Area under the ROC curve (AUC) with 95% confidence intervals (95% CI) of SARS-CoV-2 antigen measured in NPS and saliva by means of CLEIA. Patients were considered overall and after they have been subdivided on the basis of the time lapse between onset of symptoms and enrollment. Patients were classified as positive or negative on the basis of rRT-PCR on NPS.

|  | Positive<br>(N.) | Negative<br>(N.) | NPS antigen CLEIA<br>AUC<br>(95% CI) | Positive<br>(N.) | Negative<br>(N.) | Saliva antigen CLEIA<br>AUC<br>(95% CI) |
| --- | --- | --- | --- | --- | --- | --- |
| Overall | 75 | 81 | 0.939<br>(0.903-0.977) | 80 | 141 | 0.805<br>(0.740-0.870) |
| ≤ 7 days | 32 | 42 | 0.985<br>(0.965-1.00) | 39 | 94 | 0.879<br>(0.801-0.957) |
| 7-14 days | 37 | 25 | 0.897<br>(0.819-0.976) | 34 | 30 | 0.784<br>(0.668-0.899) |
| > 14 days | 6 | 14 | 0.809<br>(0.607-1.00) | 7 | 17 | 0.697<br>(0.428-0.967) |

**Table 6.**
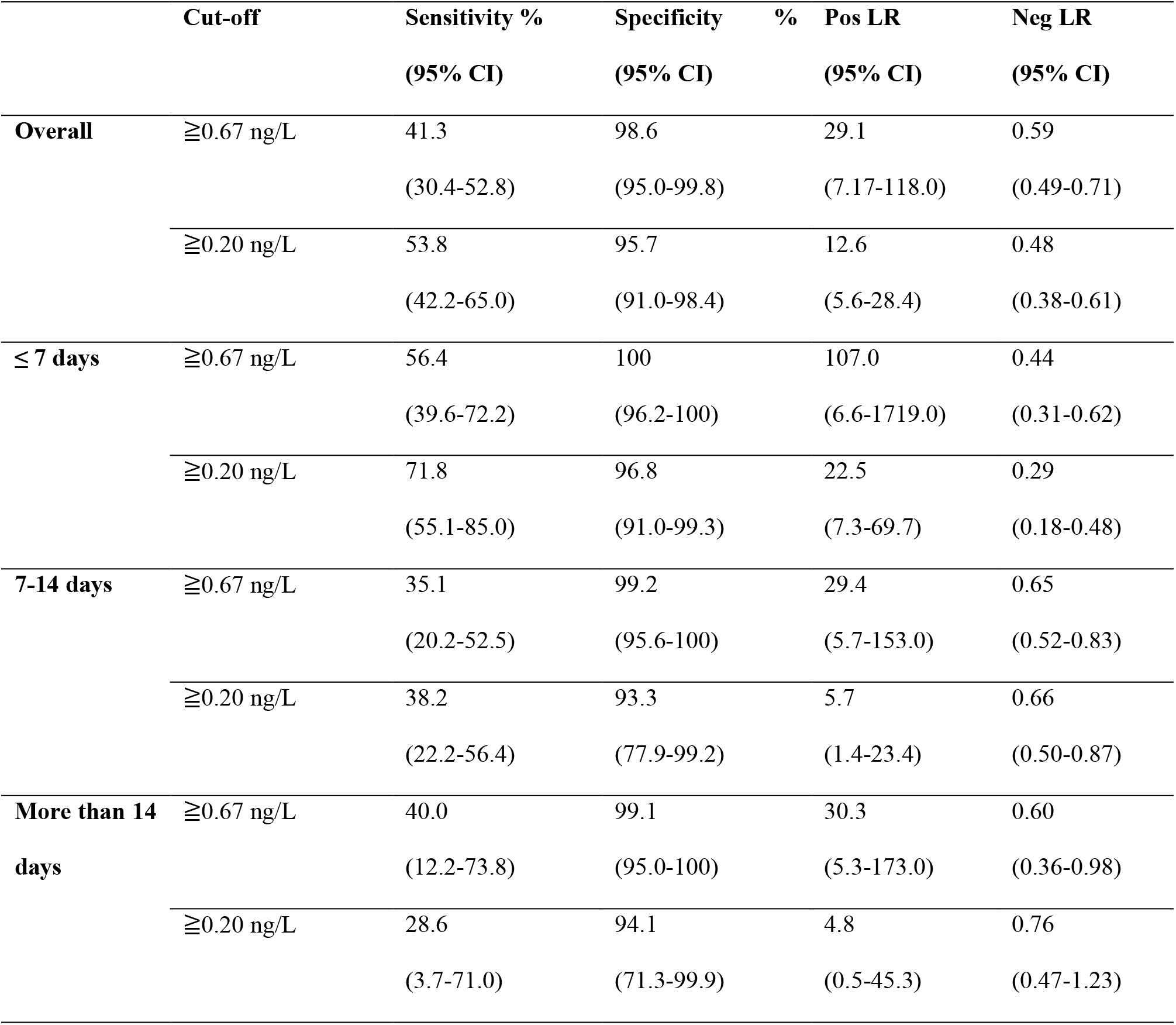
Sensitivity, specificity, positive and negative likelihood ratio (LR) with 95% confidence intervals (95% CI) of salivary CLEIA antigen. The manufacturer’s suggested cut-off (0.67 ng/L) and the manufacturer’s declared limit of quantification (0.20 ng/L) were considered as thresholds.

Figure 2 shows the individual levels of SARS-CoV-2 antigen measured in NPS (n=23) and saliva (n=32) and the corresponding Ct values obtained in a series of inpatients for whom two consecutive samples were available: one at enrollment and the other, after 7 days.

**Figure 2.**
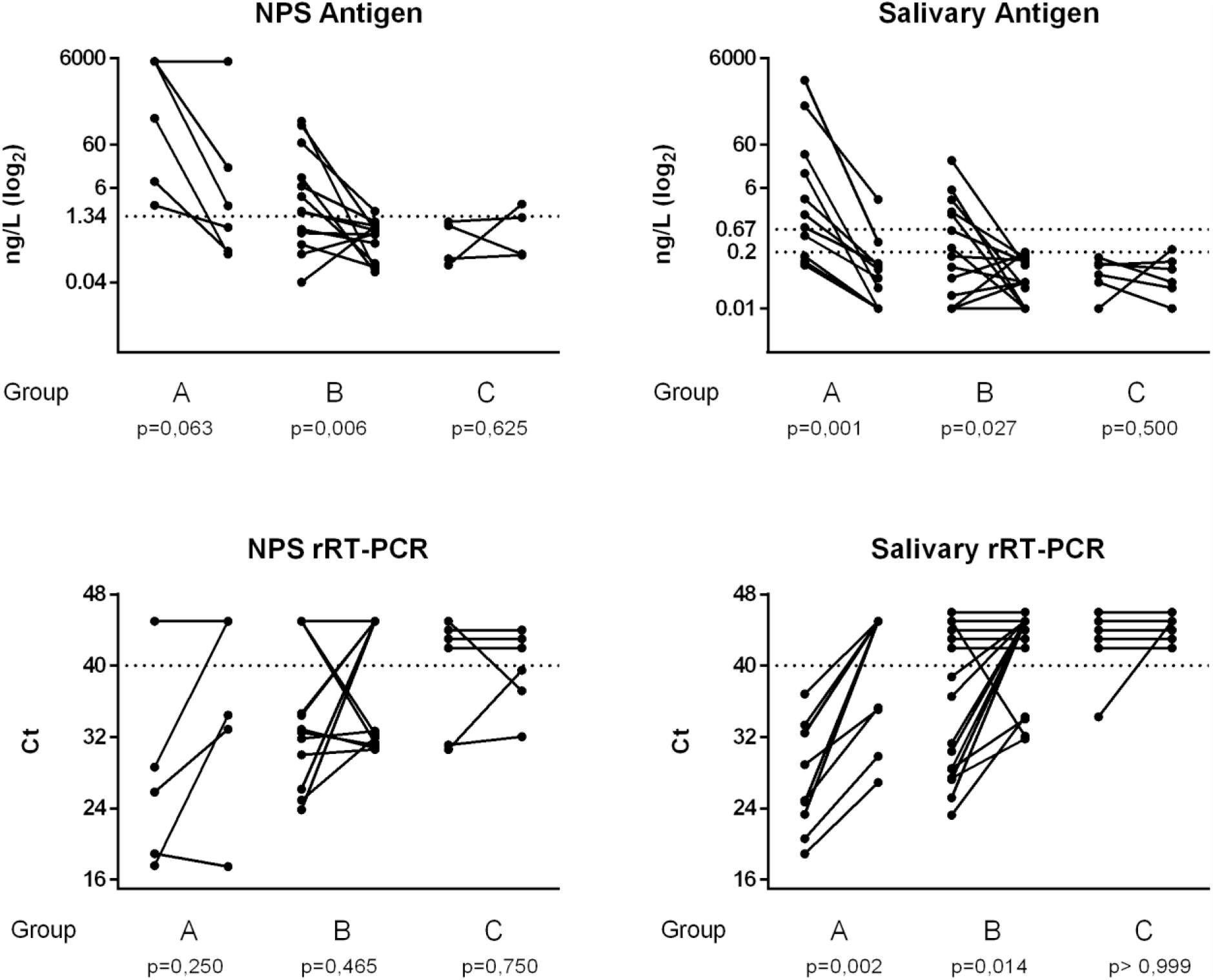
Kinetics of SARS-CoV-2 antigen and Ct values in NPS and saliva. Two consecutive samples (seven days apart) were available in a series of inpatients, who were subdivided on the basis of days from symptoms onset to enrollment in three groups: within 7 days (group A), between 7 and 14 days (group B), after 14 (group C). The p values reported were obtained after Wilcoxon rank test for paired data.

Multiple linear regression analyses were performed considering the Ct values (Orf1ab) of NPS and saliva as dependent variables, while antigen levels, clinical and demographic data as predictor variables (Table 7). The Ct vales were significantly correlated with antigen values independently from all clinical and demographic characteristics (r^2^=0.793 for NPS and r^2^=0.598 for saliva).

**Table 7.** Multiple linear regression analyses considering the Ct values (Orf1ab) of NPS and saliva as dependent variables, and the corresponding antigen levels, with clinical and demographic data as predictor variables.

|  | NPS |  |  | Saliva |  |  |
| --- | --- | --- | --- | --- | --- | --- |
|  | Coeff | 95% CI | p | Coeff | 95% CI | p |
| **Antigen (log<sub>2</sub>)** | -1.10 | -1.30 to -0.91 | <0.001 | -1.51 | -1.95 to -1.10 | <0.001 |
| **Age** | 0.02 | -0.02 to 0.06 | 0.371 | -0.01 | -0.10 to 0.08 | 0.846 |
| **Gender** | 0.27 | -1.17 to 1.70 | 0.715 | -0.81 | -4.07 to 2.44 | 0.616 |
| **Time from symptom** | 0.10 | -0.10 to 0.30 | 0.330 | 0.33 | -0.11 to 0.77 | 0.142 |
| **Fever** | 0.82 | -0.96 to 2.61 | 0.359 | -0.82 | -4.94 to 3.31 | 0.691 |
| **Dyspnea** | 0.77 | -1.21 to 2.75 | 0.440 | 1.68 | -3.03 to 6.40 | 0.476 |
| **Cough** | 0.43 | -0.93 to 1.78 | 0.532 | -0.04 | -3.10 to 3.17 | 0.981 |
| **Gastrointestinal** | -0.35 | -1.98 to 1.27 | 0.665 | 0.27 | -3.50 to 4.10 | 0.885 |
| **Pneumonia** | -0.42 | -2.50 to 1.75 | 0.703 | 3.53 | -1.32 to 8.38 | 0.150 |
| **Other** | 1.21 | -0.12 to 2.56 | 0.074 | 0.71 | -2.48 to 3.92 | 0.655 |
| **Therapy** | -0.16 | -0.9 to 0.57 | 0.656 | -0.44 | -2.12 to 1.24 | 0.603 |
| **Constant** | 30.3 | 26.4 to 34.1 | <0.001 | 28.53 | 19.97 to 37.1 | <0.001 |

Figure 3 shows the percentages of positive results for SARS-CoV-2 antigen by POC and CLEIA in NPS and saliva after subdividing samples based on viral load estimated with the Ct values at molecular analyses. POC sensitivity was satisfactory only for NPS with high viral loads (i.e. Ct values < 25), while CLEIA enabled the detection of viral particles with a good sensitivity also for Ct values above 25.

**Figure 3.**
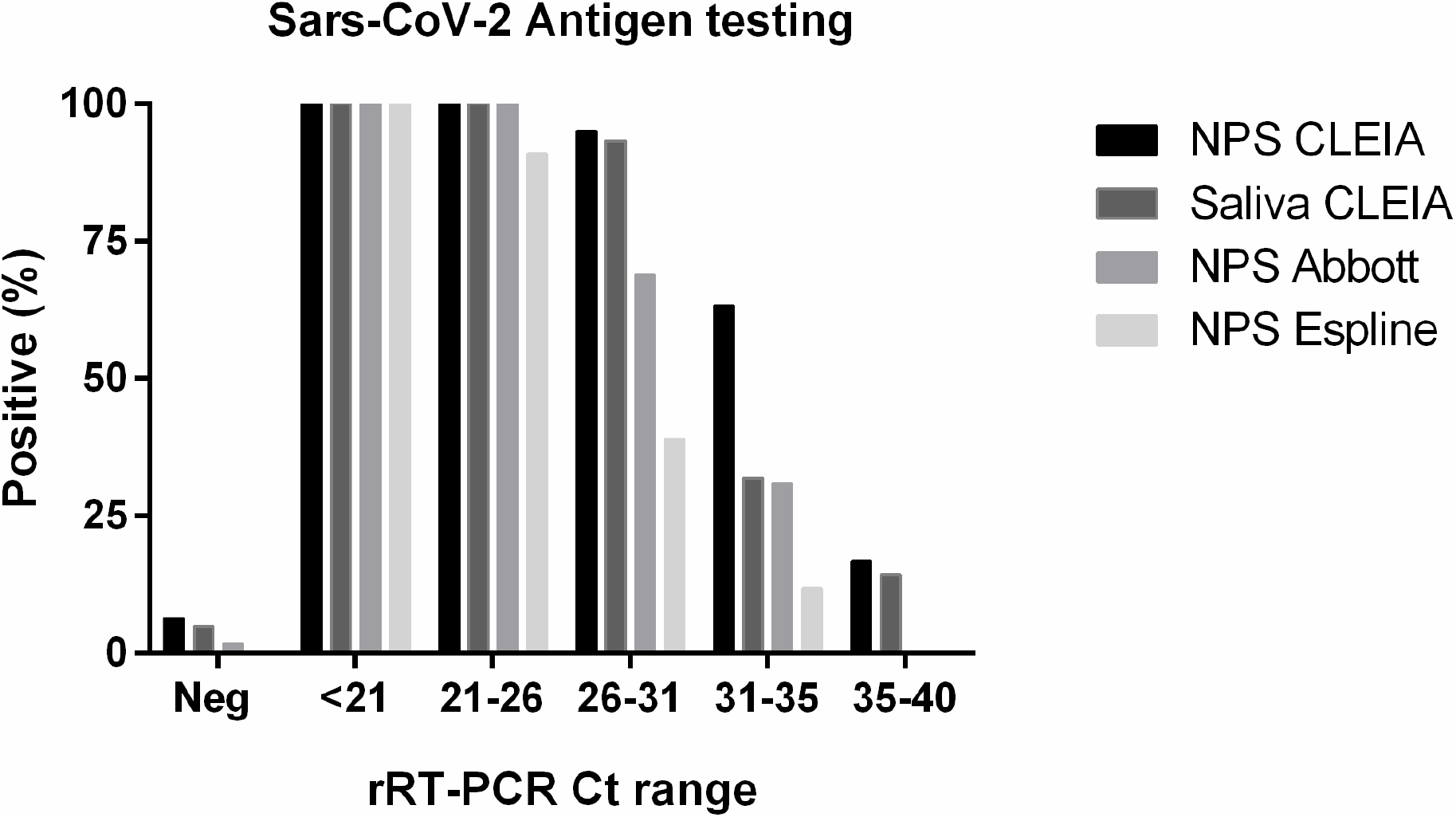
Comparison of NP and saliva SARS-CoV-2 rapid antigen testing. Percentages of positive results for SARS-CoV-2 antigen testing by means of rapid immunochromatographic assays (Abbott and Espline) and CLEIA in NPS and saliva after subdividing samples on the basis of viral load (Ct ranges) at molecular analyses.

## Discussion

There is a pressing need for novel strategies for the effective containment of a third wave of SARS-Cov-2 infection, particularly while awaiting vaccines. The early, reliable identification of SARS-CoV-2 infection appears to be the key to reducing community transmission yet the recommended diagnosis based on rRT-PCR analysis of NPS, although accurate, does not enable an early and prompt diagnosis. Moreover, it calls personnel trained in NPS collection, analysis and interpretation. SARS-CoV-2 antigen determination in NPS by point-of-care immunochromatographic assays or by chemiluminescent assays developed with laboratory instrumentation has been proposed in order to overcome these limitations, and to facilitate large scale analyses (22). This approach allows a reduction in the analytical time but does not obviate the need to perform NPS by trained personnel within dedicated medical cabinets. This bottle-neck could be overcome by using self-collected saliva. To identify the best possible strategy for detecting the infection by antigenic rapid testing, in this second wave pandemic we studied two series of subjects representing real world scenarios: inpatients with COVID-19 disease and outpatients screened for SARS-CoV-2 due to a history of positive contact or suspect clinical signs. NPS and saliva, simultaneously collected from all patients enrolled in the study, were used as matrices for antigen detection employing immunochromatographic assays and rapid CLEIA, and for viral sequences identification by rRT-PCR. As expected, the prevalence of positive rRT-PCR findings in NPS was lower among outpatients (3%) than inpatients (61%); in this latter group, it progressively declined in parallel with duration of symptoms, as the viral load, assessed by the Ct value, declined. Based on this observation, our patients’ series was subdivided according to the duration of symptoms before enrollment: less than one week, one to two weeks, and more than two weeks. The molecular detection of viral sequences in NPS and saliva gave concordant results in a high percentage of cases at the onset (92%) and in the late phases (82%) of the infection. Conversely, patients enrolled one to two weeks after symptom onset were more likely to have positive NPS results than positive saliva rRT-PCR results, in agreement with previous data in the literature (16). These findings corroborate the hypothesis that buccal mucosa and salivary glands are the first sites of viral colonization, and as saliva is the first route for viral dissemination, it represents a suitable matrix for screening asymptomatic subjects, who are known to carry a viral load comparable to that of symptomatic patients (6,23).

CLEIA antigen testing in NPS enabled a highly accurate distinction between positive and negative swabs. The area under the ROC curve was higher than 0.9, and better than that reported earlier in a Japanese series by Hirotsu et al. (20). Unlike the approach used in the cited study, we evaluated a large number of infected patients and for any patient one single sample, while Hirotsu et al. included serial samples from a limited number of infected patients. Moreover, CLEIA antigenic testing in NPS had a very high sensitivity not only in the first week, but also in the second week from symptom onset paralleling molecular findings. With respect to CLEIA, rapid immunochromatographic assays in NPS appeared very reliable for high viral loads but much less so in the presence of less abundant viral loads (i.e. Ct values ranging from 25 to 30) or even worse, for higher Ct values (>30), in agreement with data reported in the literature (19,24). This must be considered a limitation of these rapid immunochromatographic assays taking also into account that Ct values ranging from 20 to 30 are considered normal findings (25). We therefore suggest that CLEIA antigenic testing in NPS should be preferred to rapid immunochromatographic assays for obtaining accurate and fast results in the emergency setting to rapidly identify infected symptomatic patients before their hospitalization, on considering that in this context NPS collection does not represent a limitation. CLEIA antigenic testing, which requires a simple laboratory instrumentation usable by minimally trained personnel, enables the result to be obtained in 30 minutes and processes more than 100 samples per hour and can therefore be used by any emergency laboratory.

In the setting of screening asymptomatic subjects, salivary antigen might make the difference. In self-collected saliva, CLEIA antigen allowed subjects with positive to be differentiated from those with negative NPS with a good overall accuracy (0.81), which was even better in the early infection phase (accuracy 0.88) (26). The antigen levels in saliva declined more rapidly that in NPS paralleling the decline in viral load (27). Interestingly considering the patients with repeated sampling, the Ct of NPS especially in the time frame of one-two weeks from symptom onset exerted a higher variable pattern than saliva findings. This discrepancy might depend on saliva collection being more standardized than NPS collection, but also on the possibility that viral RNA fragments in the absence of infectivity could be detected in NPS by molecular testing (28). We focused our analyses mainly in this early phase because it is more representative of the possible screening scenario. By using the manufacturer’s suggested cut-off of 0.67 ng/L for CLEIA salivary antigen, in the presence of specificity close to 100%, the highest sensitivity was achieved within the first week of onset of symptoms. However, the 56.4% sensitivity observed is too low for screening programs, especially when the prevalence of disease is low, as might occur in the near future following the effects of lockdown policies. Therefore we evaluated whether an increase in sensitivity, without decreasing specificity, could be achieved by lowering the cut-off to the limit of detection suggested by the manufacturer (0.2 ng/L), and achieved 72% sensitivity and 96% specificity. We believe that CLEIA salivary testing could be applied in screening asymptomatic cohorts (e.g. in schools or farms) according to the following strategic plan: 1. Values above 0.67 ng/L to be considered positive for SARS-CoV-2; values below 0.2 ng/L to be considered negative for SARS-CoV-2; values ranging from 0.2 to 0.67 ng/L to be considered a grey zone and, for these samples, a reflex rRT-PCR activated. To be effective, this strategy should be used on a large scale, and undertaken at least once a week, in agreement with the proposal for less sensitive point of care tests with a quick return suggested as useful in surveillance when repeated at least every three days (29). However, immunochromatographic saliva testing had an unacceptable overall sensitivity of about 13%, in line with previous data obtained with the same (26), or different diagnostic systems (25); this renders it unhelpful, even if repeated daily.

The limitation of our study is represented by the low number of subjects enrolled for screening. Feasibility of saliva testing for screening programs is under evaluation at the University of Padova since October 2020, aiming to test saliva samples from about 4000 asymptomatic employees every 15 days.

In conclusion, SARS-CoV-2 antigen testing by CLEIA is fast enough to meet requirements for the early detection of the infection by any laboratory. CLEIA antigen testing in NPS might be suggested in the emergency setting, while CLEIA antigen testing with molecular reflex testing for the grey zone results in saliva is suggested by the authors of this study for large scale screening of asymptomatic subjects in risk cohorts.

## Data Availability

Data will be make available upon request

## Author’s contributions

DB and AA designed the study and drafted the manuscript. AA and NC collected the samples and clinical data. CC, FN, SM and CFZ performed the analyses. DB, AA and AP analyzed and summarized all the data. AMC and MP revised the final manuscript. All authors approved the final version of the manuscript.

## Acknowledgements

We thank all personnel of the Tropical and Infectious Disease Unit for their valuable assistance in collection of biomaterials. We thank Mrs Tamara Avellone, Cinzia Centobene, Monica Razetti and Daniela Rinaldi for their technical assistance in laboratory testing.

## List of abbreviations

(NPS): naso-pharyngeal swab
(CLEIA): chemiluminescent assay
(POC): point-of-care
(ROC): receiver operating characteristic
(AUC): area under the receiver operating characteristic curve
(SD): standard deviation.

